# Vaccine effectiveness of Pfizer-BioNTech and Oxford-AstraZeneca to prevent severe COVID-19 in Costa Rica by September and October 2021: A nationwide, observational study of hospitalisations prevalence

**DOI:** 10.1101/2021.11.08.21266087

**Authors:** Luis Rosero-Bixby

## Abstract

**Objective:** To estimate the dose-dependent effectiveness of coronavirus disease (COVID-19) vaccines to prevent severe illness in real-world conditions of Costa Rica, after the Delta variant became dominant.

**Design:** Observational study; secondary analysis of hospitalisation prevalence.

**Setting:** Nationwide adult population, Costa Rica.

**Participants:** All 3.67 million adults residents in Costa Rica by mid-2021. Public aggregated data of 5978 hospital records from 14th September to 20th October, 2021 and 6.1 million vaccination doses administered.

**Interventions:** Vaccination with Pfizer-BioNTech (78%) and Oxford-AstraZeneca (22%).

**Main outcome measures:** Prevalence of COVID-19-related hospitalisations

**Results:** Vaccine effectiveness to prevent hospitalisation (VEH) was estimated as 93.4% (95% confidence interval [CI]: 93.0 to 93.9) for complete vaccination and 76.7% (CI: 75.0 to 78.3) for single-dose vaccination among adults of all ages. VEH was lower and more uncertain among older adults aged 58 years and above: 92% (CI: 91% to 93%) for those who had received full vaccination and 64% (CI: 58% to 69%) for those who had received partial vaccination. Single-dose VEH declined over time during the study period, especially in the older age group. Estimates were sensitive to possible errors in the population count used to determine the residual number of unvaccinated people in groups with high vaccine coverage.

**Conclusion:** The Costa Rican vaccination programme that administered Pfizer and Oxford vaccines are highly effective to prevent COVID-19-related hospitalisations after the Delta variant had become dominant. Moreover, a single dose is reasonably effective, justifying the continuation of the national policy of postponing the application for the second dose of the Pfizer vaccine to accelerate the vaccination and increase the number of people being vaccinated. Timely monitoring of vaccine effectiveness is important to detect eventual failures and motivate the public based on information that the vaccinations are effective.

**Summary Box:** *What is already known on the topic:*

- The Costa Rican Social Security Fund provides vaccinations in Costa Rica, and they use Pfizer-BioNTech and Oxford-AstraZeneca for the vaccinations. The Delta variant became predominant in Costa Rica by September 2021
- Real-world estimates of these vaccines effectiveness to prevent hospitalisations range from 90% to 98% for two doses and from 70% to 91% for a single dose. Almost all of these estimates predate the Delta variant.
- There are controversies regarding the effectiveness of a single dose of COVID-19 vaccine after the emergence of the Delta variant.

*What this study adds:*

- Vaccine effectiveness to prevent hospitalisation as estimated as 93.4% (95% CI: 93.0 to 93.9) for complete vaccination and 76.7% (CI: 75.0 to 78.3) for single-dose vaccination among adults of all ages by October 2021.
- These study findings suggest that a single dose of COVID-19 vaccination is reasonably effective to prevent hospitalisations.
- Therefore, the application for the second dose of the Pfizer vaccine can be postponed beyond the three weeks recommended by the fabricant to accelerate vaccination coverage.

## Introduction

Controversies regarding the possible lack of effectiveness of a single dose of coronavirus disease (COVID-19) vaccination arose with the emergence of the Delta variant, a more contagious variant of COVID-19. In Costa Rica, the COVID-19 cases caused by the Delta variant increased from 11% in a sample of new infections in the last week of June 2021 to 55% in the first week of August and to 100% in the last week of September 2021.(1) In the same period, the incidence of COVID-19 increased from 288 to 445 daily cases per million population, despite the rapid increase in the proportion of vaccinated people from 32% on 28th June to 66% on 27th September 2021.(2)

The universal public health care system of Costa Rica, which is provided by the Costa Rican Social Security Fund (CCSS, acronym in the Spanish language), has been the single source of COVID-19 vaccination in the country. The CCSS uses two vaccines, the mRNA from Pfizer-BioNTech and the adenovirus vector from Oxford-AstraZeneca (i.e., Pfizer and Oxford vaccines). By 20th October 2021, 85% of the 3.7 million adult residents had been vaccinated with at least one dose and 59% with two doses.(3) Older adults aged 58 years and above received only the Pfizer vaccine; 87% received the two doses in a 3-week interval and 13% in an 8-to 12-week interval. Among the adults below 58 years of age, 77% received the Pfizer and 23% received the Oxford vaccines, mostly in an 8-to 12-week interval between doses. The Costa Rican vaccination program had a sttrong initial focus on older adults (aged 58 years and older, as defined by the government). By 1st June 2021 more than 80% of the population in this age group had been vaccinated compared to 17% in the 40 to 57 years age group and 7% in the 20 to 39 years age group. Due to this initial focus on older adults, the proportion of older adults with more than a 6-month period after their second dose was growing rapidly during the study period from 2% on 14th September to 24% on 20th October 2021.

At the time of this study, the literature reported the following real-world estimates (based on observational studies rather than randomised trials) of COVID-19 vaccine effectiveness to prevent hospitalisations (VEH). The two-dose Pfizer VEH was 97% in Israel,(4) 98% in Ontario, Canada in a vaccination program with an allocated 77% Pfizer vaccine,(5) 91% in the United States in the first 4 months after full vaccination,(6) 90% in California when 93% of the COVID-19 was caused by the Delta variant,(7) and 90% in Qatar according to a preprint study on only Delta variant cases.(8) The near 100% VEH estimates in Israel and Ontario were obtained before the emergence of the Delta variant.

Single-dose VEH estimates were 70% in the Ontario vaccination program, which was mostly Pfizer vaccination,(5) 80% in England for both Pfizer and Oxford vaccines,(9) and 91% and 88% in Scotland for Pfizer and Oxford vaccines, respectively.(10) All these estimates predated the Delta variant.

The objective of this study was to estimate the dose-dependent effectiveness of coronavirus disease (COVID-19) vaccines to prevent severe cases of COVID-19, as measured by the prevalence of hospitalisations, in a middle-income country, Costa Rica. These estimates were based on secondary analysis of COVID-19 related hospitalized individuals from 14th September to 20th October 2021.

A central motivation for this study was to assess the rationale of the national policy of increasing the interval between the first and second doses of the Pfizer-BioNTech vaccine from 3 to 12 weeks. The policy was adopted on 1 June 2021. If the data show that the effectiveness of a single dose of the vaccine is weak, this policy must be revised and the interval between doses should be reduced as much as possible.

## Methods

This observational, nationwide study used a cross-sectional prevalence design. The study performed secondary analysis of official statistics and reports. It compared the COVID-19 related hospitalisation prevalence among the unvaccinated population with the prevalence among the semi-vaccinated and fully vaccinated populations at six time points, 1 week apart, from 14th September to 20th October 2021.

The VEH estimates used three sources of data:

1. A series of weekly reports presented by the Department of Health Statistics to the Board of Directors of the CCSS,(11) which is the most important information. These reports show the distribution by vaccination status of COVID-19 related hospitalisations by the ages of patients. CCSS officers linked the databases of hospitalisations and vaccinations to determine the vaccination status of hospitalised individuals and some demographic characteristics such as age and sex. For 2% of the hospital records, the vaccination status was not established.
2. The time series of the number of first and second doses of the COVID-19 vaccines administered (6.1 million until October 20, 2021) according to population age groups as reported weekly by the CCSS.(3) I used these data to estimate the nationwide populations of semi-and fully vaccinated individuals by age groups in the six time points of the study. No adjustments were made for changes in demographics (no vaccinated individual died, out-migrated, or changed age bracket) in these populations since the impact of these changes are minuscule considering the short study period.
3. The mid-2021 nationwide population estimate by the National Institute of Statistics and Censuses (INEC).(12) I used this estimate to determine the residual group of unvaccinated individuals, who represented the control group in the analysis. It was assumed that there were no changes in the population from the date of the estimate to the study dates.

The outcome variable was “being hospitalised due to COVID-19.”

The intervention variables were the two vaccination schemes as defined by the CCSS:

1. Partially or *semi-vaccinated* individuals: 15 or more days after the first dose and either less than 15 days after the second dose or no second dose.
2. *Fully vaccinated* individuals: 15 or more days since the second dose.

### Statistical methods

All analyses were stratified according to three age groups: 20–39, 40–57, and 58 years or more, which are groups defined in the priority calendar of the national vaccination program.

VEH was estimated as one minus the hospitalisation-prevalence ratio of vaccinated to unvaccinated populations. Given the strong confounding effects of age, the Mantel-Haenzel technique was used to aggregate the age-specific estimates into a summary indicator for the entire adult population.(13) No imputations were made for 2% of hospitalisations that had missing data, which were assumed to be randomly distributed. Estimates were obtained using the Stata-17 statistical software, “epitab” commands.(14)

While the number of vaccinated persons is a direct count of administered vaccines, the number of unvaccinated persons was an indirect estimate as the residual: population minus the number of vaccinated people. Error in the population estimate could amplify in the residual number of the unvaccinated individuals. A sensitivity analysis was performed to assess the impact of this potential error on the VEH.

### Patient and public involvement

Interactions of the author with journalists, health professionals, and lay individuals suggested that they had difficulties interpreting the data provided by health authorities regarding the vaccination status of COVID-19 hospitalisations. For example, the official report of a substantial number of hospitalized people who were fully vaccinated, some interpreted as proof that the COVID-19 vaccines were not effective. This study is a response to this wrong interpretation.

## Results

Overall, the study included data of 3.67 million individuals, the entire adult population of Costa Rica. Of this population, 47% were in the younger group, 31% in the intermediate group, and 22% in the older group. The number of hospital records assessed in the six time periods was 5978, excluding 138 records with missing information. The Appendix table shows the data used in the study, namely the number of participants (the population) and COVID-19-related hospitalisations. The table also shows the resulting prevalence proportions as well as the estimated VEH.

The highest rates of hospitalisation occurred among older unvaccinated individuals, with a prevalence of 3537 to 4754 per million people. The lowest rates of hospitalisation occurred among fully vaccinated younger adults, with prevalence ranging from 5 to 11 per million people, approximately 400 times lower than the unvaccinated old. Hospitalisation prevalence increased with age and was substantially higher among unvaccinated individuals. Over time, the prevalence proportions reflected the fact that the incidence of the pandemic in Costa Rican had reached its peak at the beginning of September, followed by peaks of hospitalisations after 2 weeks.(15)

As stated previously, VEH was estimated by comparing the prevalence of COVID-19 hospitalisations in the partially or fully vaccinated group to that in the unvaccinated group. Figure 1 shows all VEH estimates with 95% confidence intervals.

**Fig 1.**
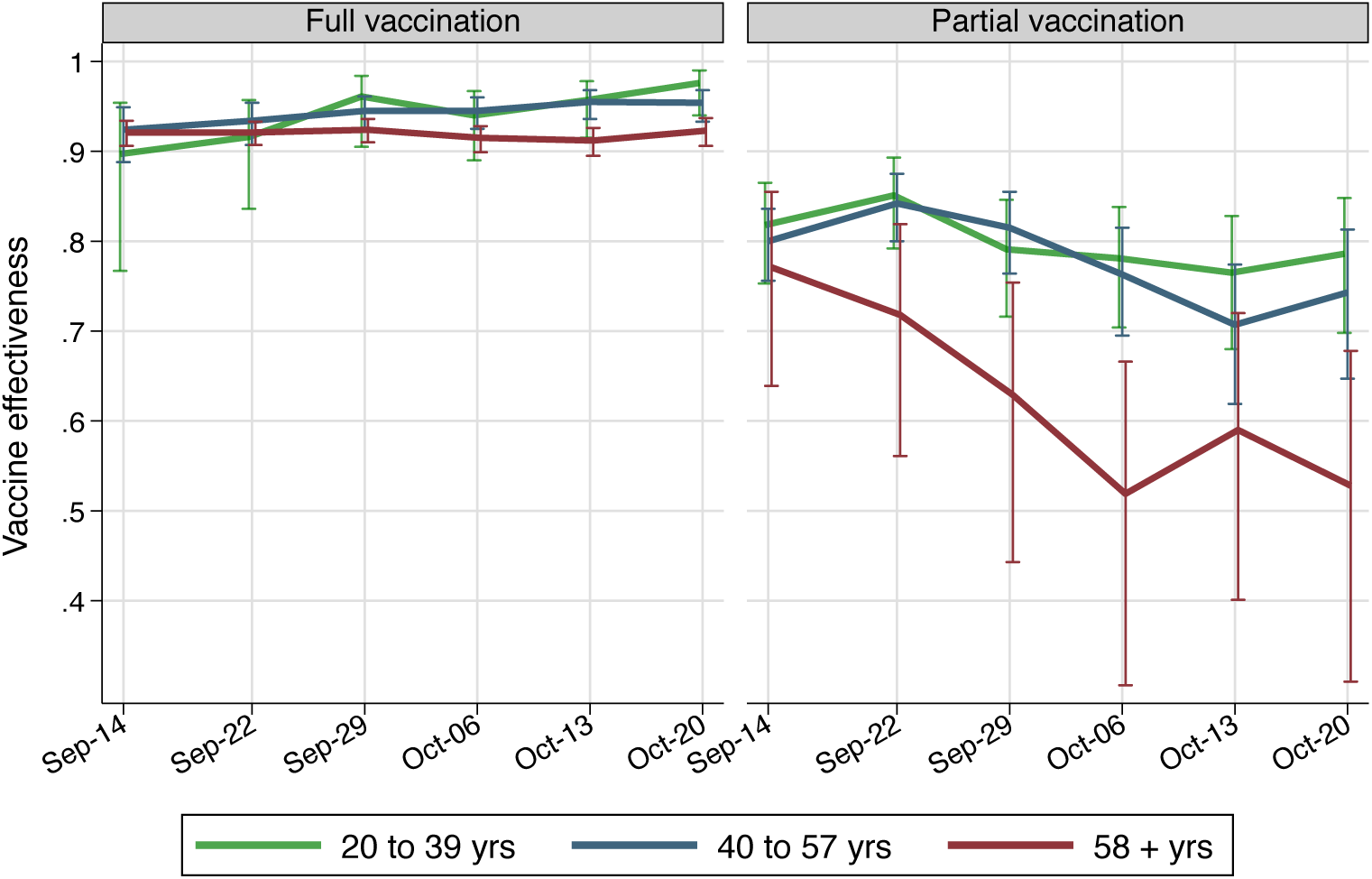
COVID-19 VEH estimates and their 95% confidence intervals by age groups and cross-section dates. Costa Rica, September and October 2021.

The VEH for full vaccination ranges between 0.90 and 0.98 in the three age groups and six time points, with a statistically significant ascending time trend in the youngest group. In contrast, the VEH for partial vaccination significantly declined during the study period, especially in the older age group. The average weekly decline is 0.01, 0.02, and 0.05 in the three age groups, respectively. The range of partial vaccination effectiveness estimates ranges from 0.52 to 0.77 among older adults and from 0.71 to 0.84 among the other adult groups. Estimates of VEH for partial vaccination are substantially less precise than that for full vaccination, as shown by the wider confidence interval, especially in the older-adult group.

Table 1 shows the summary indicators of VEH obtained after pooling the data from the six observed time periods. These estimates represent the status of the COVID-19 vaccination in early October 2021 in Costa Rica. The age-adjusted estimates for all adults suggest a VEH of 93.4% (CI: 93.0 to 93.9) for the full vaccination scheme of two doses and 77% (CI: 75.0 to 78.3) for partial vaccination with a single dose. Older adults showed slightly lower VEH for full vaccination (92%, CI: 91.4 to 92.5) and substantially lower VEH for partial vaccination (64%, CI: 57.5 to 69.4) compared to the other age groups. Majority of the COVID-19 hospitalisation were probably caused by the Delta variant infection, which was dominant at the time of the study according to the Costa Rican genomic tracking system of the variants of concern.(1)

**Table 1.** VEH for full and partial COVID-19 vaccination by age groups and relative risks (RR) of the unvaccinated. Costa Rica, October 2021.

| Age groups | Full vaccination |  | Partial vaccination |  |
| --- | --- | --- | --- | --- |
|  | VEH/RR | (95% Conf. Int.) | VEH/RR | (95% Conf. Int.) |
| 20 - 39 | 0.949 | (0.932 to 0.963) | 0.801 | (0.774 to 0.825) |
| 39 - 57 | 0.947 | (0.939 to 0.954) | 0.792 | (0.770 to 0.811) |
| 58 + | 0.920 | (0.914 to 0.925) | 0.639 | (0.575 to 0.694) |
| *All 20+* |  |  |  |  |
| Crude | 0.806 | (0.795 to 0.816) | 0.835 | (0.824 to 0.846) |
| Age adjusted* | 0.934 | (0.930 to 0.939) | 0.767 | (0.750 to 0.783) |
| Relative risk of the unvaccinated, compared to the: |  |  |  |  |
|  | Fully vaccinated |  | Partially vaccinated |  |
| 20 - 39 | 19.8 | (14.7 to 26.7) | 5.0 | (4.4 to 5.7) |
| 39 - 57 | 18.9 | (16.4 to 21.7) | 4.8 | (4.3 to 5.3) |
| 58 + | 12.5 | (11.6 to 13.4) | 2.8 | (2.4 to 3.3) |
| *All 20+* |  |  |  |  |
| Crude | 5.1 | (4.9 to 5.4) | 6.1 | (5.7 to 6.5) |
| Age adjusted* | 15.3 | (14.2 to 16.4) | 4.3 | (4.0 to 4.6) |

\* Mantel-Haenzel estimate

Another metric for demonstrating vaccine effectiveness is by comparing the risk of being bedridden between the unvaccinated and vaccinated, as reported in the second panel of Table 1. These metrics may be more meaningful for lay people. Table 1 shows that risk of hospitalisation in the unvaccinated was 15 (CI: 14 to 16) times higher than that in the fully vaccinated and 4.3 (CI 4.0 to 4.6) times higher than that in the partially vaccinated.

There were significant differences between the crude and age-adjusted estimates for the entire adult population (Table 1), Age was a significant confounder in these data. Older individuals had an extremely higher risk of being hospitalised and were more likely to be fully vaccinated than that had by the other adults. These two associations make that for the all-age estimate, the crude VEH were substantially lower in the older adults than in the other age groups and, thus, lower than its real magnitude, which was estimated by the simple method proposed by Mantel and Haenzel in 1959 as a weighted average of age-specific figures. The crude all-age VEH for full vaccination was 81% compared to the 93% when the age was adjusted.

Figure 2 summarises the results of the sensitivity analysis to possible errors in the population data input. Errors of plus or minus 1% in the population input would bias the VEH estimates by less than 0.01, except in partially vaccinated older adults, where a change of 0.05 would occur. Larger errors of plus or minus 5% in the population input would alter the VEH by 0.02 in the two younger groups and would strongly bias the estimate between 0.04 and 0.27 for partial vaccinations. VEH estimates for older adults appear especially sensitive to errors in the population data, which originates from the very high vaccination coverage reached by this age group: approximately 91.5% fully and 2% partially vaccinated at the end of the study period. Small errors in population inputs substantially amplify the residual estimates of unvaccinated individuals when vaccine coverage is high.

**Fig 2.**
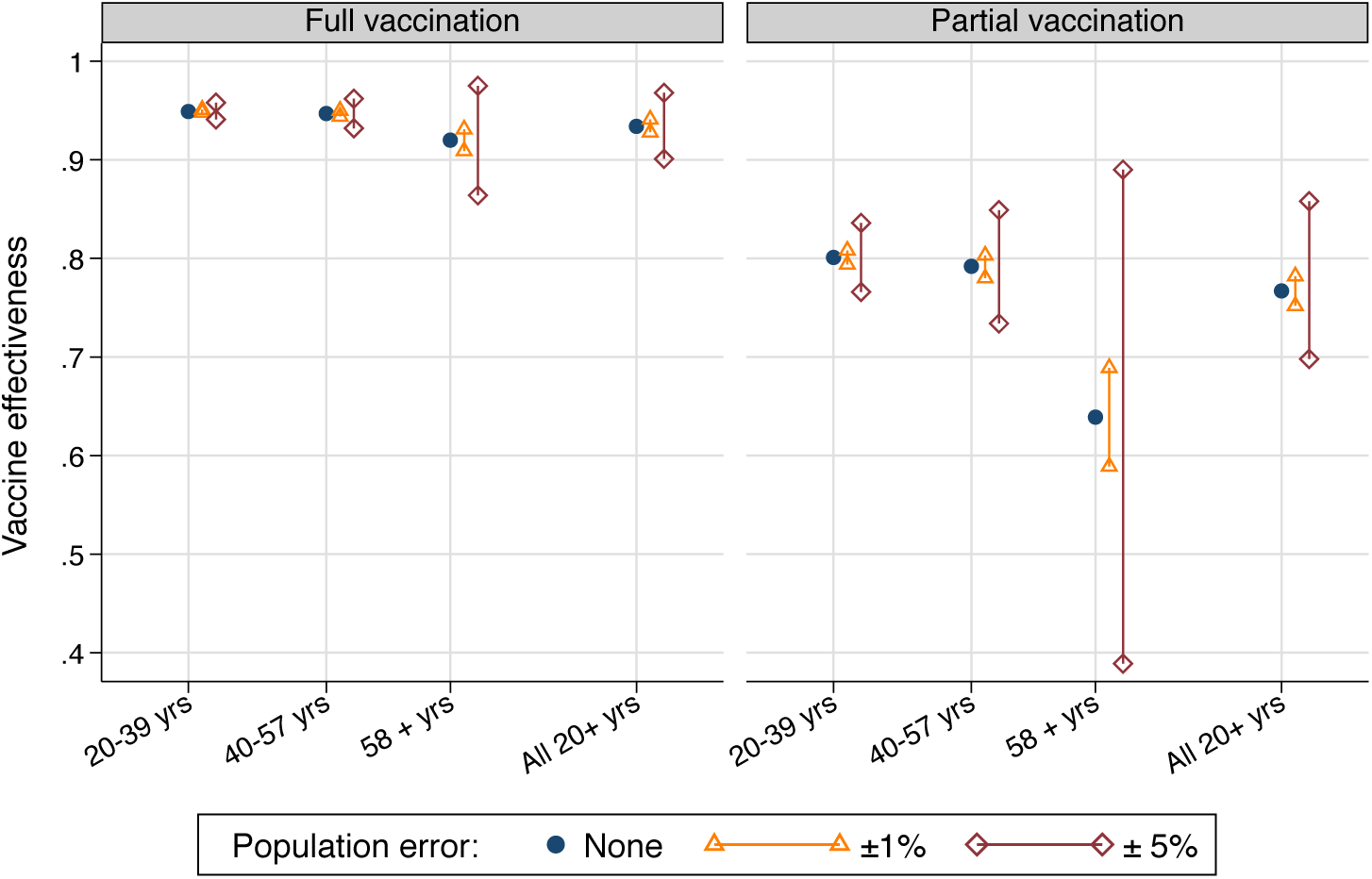
Sensitivity of VEH estimates to possible errors in the population number used as input.

## Discussion

COVID-19 vaccine effectiveness to prevent severe illness, as identified by the prevalence of hospitalisations, is 93% for the complete vaccination scheme of two doses in the adult population of Costa Rica. Among the subgroup of older adults aged 58 years and older, VEH was slightly lower (92%) than that in the other subgroups. The corresponding VEH estimates for a single vaccine dose were 77% for all adults and 64% for the older subgroup. Costa Rica uses two COVID-19 vaccines, Pfizer-BioNTech (78% of the vaccines administered) and Oxford-AstraZeneca (22% of the vaccines administered).(3) The estimates in this article largely reflect vaccine effectiveness against the Delta variant since it was the dominant variant in the country during the study period.(1)

The high VEH for the single-dose showed a statistically significant decline in time trend by 0.01 or 0.02 per week in adults aged 20–57 years and 0.05 per week in the older age group. It is important to note that the decline did not occur in the VEH for full vaccination. If it was caused by the spread of the Delta variant or by depletion of the immunity provided by the vaccine, the decline should have also occurred among the fully vaccinated. A plausible explanation is that the partially vaccinated may have comprised two types of people, 1) those who are in the group temporarily while waiting for the second shot and 2) those who intentionally avoid the second shot. The VEH decline might reflect an increased share of the second subgroup as coverage approaches 100%.

This study’s estimate of 93% VEH for two doses of the Pfizer and Oxford vaccines is within the 90% to 98% range of previous real-world estimates obtained in Israel, Ontario, the United States, California, and Qatar.(4) (5) (6) (7) (8) Only the studies performed in California and Qatar reported proportions that had majority of the Delta variant infections, as reported in the present study.

The 77% single-dose VEH for a single dose of the vaccines is also within the range from 70% to 91% of studies performed in Ontario, England, and Scotland.(5, 9) (10) However, these previous estimates predated the Delta variant. This Costa Rican estimate is thus the first study that has reported that the Delta variant did not substantially reduce the effectiveness of a single dose of Pfizer or Oxford vaccines to prevent hospitalisation.

Strengths of this observational study are that it shows the effectiveness of COVID-19 vaccines in the real-world conditions of a middle-income country, which is outside the hyper-controlled conditions of clinical trials, their effectiveness after the Delta variant become dominant, and provision of a dose-specific effectiveness of the vaccines.

The secondary analysis of existing aggregated data in this study is an inexpensive design that can be broadly used to produce quick estimates for timely monitoring of vaccine effectiveness.

Being a nationwide study, it is free of issues regarding sampling bias and random errors derived from small sample sizes.

The outcome variable used in this study, being hospitalised due to COVID-19 related conditions, is a definite count that is mostly free of classification errors. A threat to its validity as a measure of severe COVID-19 infections could occur if proportions of the population have poor access to hospital care. However, this is not the case in the universal health care system of Costa Rica.

The statistics of dose-specific vaccinated people are probably accurate since the sole provider of vaccines in the country digitally records on real-time information of every single vaccine shot administered. The database for this information is also used for inventory control purposes and for providing digital vaccination certificates to the population. If there were widespread errors, they would be certainly noticed by these other uses of the data.

This observational study has the well-known limitations of non-randomised, non-blinded trials including selection biases, such as the early vaccination of older people in Costa Rica (which biased the crude VEH estimates, as shown in this study), and other confounders such as the risk-taking behaviour modification of some individuals after vaccination. The VEH estimates in this study should be interpreted as associations between vaccination status and hospitalisation rather than the true causal effects of vaccination.

A more specific limitation of the method used in this study is that it requires high-quality data of the population count to obtain a valid estimation of the number of unvaccinated individuals. Errors in the population count affects the number of people who have not been vaccinated, especially as the vaccination coverage approaches 100%, as is the case for the older adults in Costa Rica. However, it must be noted that Costa Rica is considered to have accurate demographic data.(16)

The lack of specific results for each brand of vaccines used in Costa Rica, as well as the lack of estimates of the vaccine effectiveness to prevent COVID-19, may also be limitations to the interpretation of these study results.

### Conclusions and policy implications

The Costa Rican vaccination programme, based on the Pfizer and Oxford vaccines, is highly effective to prevent COVID-19-related hospitalisations even after the Delta variant had become dominant. Completing the two-dose scheme clearly provides more protection than that by the single dose, and this result must always be the goal of vaccination policies.

However, since a single dose showed that maintained reasonable effectiveness against the Delta variant, the continuation of the national policy that the second doses of the Pfizer vaccine should be postponed for few weeks may be granted.

Timely monitoring of vaccine effectiveness appears feasible with procedures that are analogous to those used in this study. It is important to continue the monitoring of vaccine effectiveness to detect eventual failures in the vaccination program and motivate the public by showing that vaccinations are having an impact.

## Data Availability

All data produced in the present work are contained in the manuscript (Appendix table)

## Acknowledments

the author thanks Susana López Delgado, Statistician of the CCSS, for providing the reports with the aggregated data on covid-19 related hospitalizations by vaccination status.

---

**Appendix table.** Data used to estimate Covid-19 related hospitalisation prevalence and vaccine effectiveness (VEH) by vaccination status and age at six points in time. Costa Rica.

| Date and Vaccination status | Age 20 to 39 years |  |  |  | Age 40 to 57 years |  |  |  | Age 58 years or more |  |  |  |
| --- | --- | --- | --- | --- | --- | --- | --- | --- | --- | --- | --- | --- |
|  | Population | C-19 hospitalised | Prevalence per million | VEH | Population | C-19 hospitalised | Prevalence per million | VEH | Population | C-19 hospitalised | Prevalence per million | VEH |
| **14-Sep-2021** |  |  |  |  |  |  |  |  |  |  |  |  |
| Unvaccinated | 602,453 | 189 | 314 |  | 243,809 | 302 | 1,239 |  | 63,883 | 302 | 4,226 |  |
| Semi vaccinated | 944,645 | 54 | 57 | 0.82 | 593,039 | 147 | 248 | 0.80 | 20,660 | 147 | 968 | 0.77 |
| Fully vaccinated | 185,102 | 6 | 32 | 0.90 | 287,252 | 27 | 94 | 0.92 | 732,857 | 27 | 333 | 0.92 |
| **22-Sep-2021** |  |  |  |  |  |  |  |  |  |  |  |  |
| Unvaccinated | 596,169 | 204 | 342 |  | 240,656 | 302 | 1,255 |  | 63,374 | 302 | 4,560 |  |
| Semi vaccinated | 824,275 | 42 | 51 | 0.85 | 458,855 | 91 | 198 | 0.84 | 16,322 | 91 | 1,287 | 0.72 |
| Fully vaccinated | 311,756 | 9 | 29 | 0.92 | 424,589 | 35 | 82 | 0.93 | 737,704 | 35 | 361 | 0.92 |
| **29-Sep-2021** |  |  |  |  |  |  |  |  |  |  |  |  |
| Unvaccinated | 551,141 | 180 | 327 |  | 224,643 | 287 | 1,278 |  | 62,052 | 287 | 4,754 |  |
| Semi vaccinated | 790,324 | 54 | 68 | 0.79 | 355,344 | 84 | 236 | 0.81 | 14,192 | 84 | 1,762 | 0.63 |
| Fully vaccinated | 390,735 | 5 | 13 | 0.96 | 544,113 | 38 | 70 | 0.95 | 741,156 | 38 | 362 | 0.92 |
| **6-Oct-2021** |  |  |  |  |  |  |  |  |  |  |  |  |
| Unvaccinated | 469,731 | 165 | 351 |  | 195,425 | 250 | 1,279 |  | 58,864 | 250 | 4,485 |  |
| Semi vaccinated | 740,414 | 57 | 77 | 0.78 | 263,446 | 80 | 304 | 0.76 | 14,829 | 80 | 2,158 | 0.52 |
| Fully vaccinated | 522,055 | 11 | 21 | 0.94 | 665,229 | 47 | 71 | 0.94 | 743,707 | 47 | 382 | 0.91 |
| **13-Oct-2021** |  |  |  |  |  |  |  |  |  |  |  |  |
| Unvaccinated | 387,924 | 130 | 335 |  | 164,113 | 194 | 1,182 |  | 53,971 | 194 | 4,150 |  |
| Semi vaccinated | 724,720 | 57 | 79 | 0.77 | 227,889 | 79 | 347 | 0.71 | 17,645 | 79 | 1,700 | 0.59 |
| Fully vaccinated | 619,556 | 9 | 15 | 0.96 | 732,098 | 39 | 53 | 0.95 | 745,784 | 39 | 365 | 0.91 |
| **20-Oct-2021** |  |  |  |  |  |  |  |  |  |  |  |  |
| Unvaccinated | 354,165 | 108 | 305 |  | 150,857 | 146 | 968 |  | 51,176 | 181 | 3,537 |  |
| Semi vaccinated | 704,530 | 46 | 65 | 0.79 | 208,980 | 52 | 249 | 0.74 | 18,588 | 31 | 1,668 | 0.53 |
| Fully vaccinated | 673,505 | 5 | 7 | 0.98 | 764,263 | 34 | 44 | 0.95 | 747,636 | 204 | 273 | 0.92 |
| **Pooled (averages)** |  |  |  |  |  |  |  |  |  |  |  |  |
| Unvaccinated | 493,597 | 163 | 329 |  | 203,251 | 247 | 1,200 |  | 58,887 | 253 | 4,285 |  |
| Semi vaccinated | 788,151 | 52 | 66 | 0.80 | 351,259 | 89 | 264 | 0.78 | 17,039 | 85 | 1,590 | 0.63 |
| Fully vaccinated | 450,452 | 8 | 20 | 0.94 | 569,591 | 37 | 69 | 0.94 | 741,474 | 65 | 346 | 0.92 |

